# Adult-Learning Newborn Medicine Curriculum Improves Knowledge in a Low-Resource Neonatal Unit in Sierra Leone

**DOI:** 10.64898/2026.06.02.26354766

**Authors:** McGeofrey Mvula, Amee Amin, Monika S Patil, Gregory Valentine, Beatha Mukarwego, Sheron Wagner, Isata Dumbuya, Lily Lou, Usman Sanni, Anne R Hansen

**Affiliations:** Koidu Government Hospital, Partners in Health Sierra Leone, Kono District, Sierra Leone; Neonatology, Pediatrix Medical Group, Houston, Texas, United States of America; Department of Pediatrics, Division of Neonatology, Baylor College of Medicine, Houston, Texas, United States of America; Division of Neonatology, University of Washington/Seattle Children’s Hospital, Seattle, Washington, United States of America; Louise Herrington School of Nursing, Baylor University, Dallas, Texas, United States of America; Department of Pediatrics, University of Illinois Health, Chicago, Illinois, United States of America; Department of Pediatrics, Harvard Medical School, Boston, Massachusetts, United States of America

## Abstract

**Background:** Sierra Leone’s neonatal mortality rate is among the highest in the world. Koidu Government Hospital opened a Special Care Baby Unit (SCBU) in 2020. To increase knowledge of the SCBU health care providers (HCPs), a neonatal curriculum was implemented to facilitate HCP education on management of neonatal conditions. The aim of this study was to understand the effect of the curriculum on knowledge acquisition and the perception of the teaching methodologies among participating HCPs.

**Methods:** US-based mentors facilitated a two-phase, flipped classroom, virtual neonatal medicine curriculum between October 2024 and April 2025, followed by one-week in-person education sessions with SCBU HCPs. With each phase, participants completed pre- and post-test educational assessments. At the end of the curriculum, they completed a subjective assessment to capture perceptions related to the quality of teaching methodologies integrated within the curriculum. Wilcoxon signed rank test was used to assess pre-versus post-test change. Descriptive statistics were used to analyse the subjective assessment.

**Results:** Thirty-eight participants completed the educational assessments, 30 (79%) took all four pre- and post-tests; 25/38 (65.8%) were female, 27 (71.1%) were nurses. Median correct answers for both phases increased from the pre-to post-test for individual learners [Phase 1, pre-test 14/27 (51.9%), post-test 23/27 (85.2%), p<0.001], [Phase 2, pre-test 14/25 (56.0%), post-test 23/25 (92.0%), p <0.001]. Thirty-one participants completed the subjective assessment, of whom 96.8% (30/31) rated the curriculum to be “very effective.” All 31 participants indicated that the in-person instruction was “very helpful.” Through open text responses, they offered valuable insight into challenges, strengths, and next steps.

**Conclusion:** This neonatal curriculum resulted in significantly increased knowledge and was well regarded. Adapting this curriculum or similar curricula show promise to improve the quality of care for small and/or sick neonates in low resource settings.

## Background

Although substantial progress has been made in reducing the neonatal mortality rate (NMR) in response to the Millenium Development Goals, the rate of improvement must be accelerated to achieve the Sustainable Development Goal (SDG) of ≤ 12 deaths/1,000 live births.^1^ Sierra Leone’s NMR, at 29.6/ 1000 live births, is higher than the average for Sub-Saharan Africa and considered one of the highest in the world.^2^ The World Health Organization (WHO) calls for skilled, competent health care providers (HCPs) for each level of neonatal illness^3^

Neonatal care services in Sierra Leone are concentrated mostly in the capital city of Freetown where only 15% of the country’s population lives.^4^ To address the resulting lack of accessibility to neonatal services for the remainder of the population, Sierra Leone’s Ministry of Health established Special Care Baby Units (SCBUs) in district hospitals throughout the country. The HCPs who work in SCBUs include nurses, midwives, non-specialty-trained doctors (medical officers [MOs]), and community health officers (CHOs).^5^ District-level SCBUs offer multiple benefits including enabling HCPs to develop specialized knowledge and experience, and removing sick neonates from crowded paediatric wards limiting exposure to other patients with community acquired infections. The success of SCBUs hinges largely on the HCPs level of training in neonatal care.

Koidu Government Hospital (KGH), with support from Partners In Health Sierra Leone, opened a SCBU in 2020, to improve care for small and/or sick neonates (SSNBs). The SCBU staff requested advanced training in neonatal medicine.

The need for innovative and effective strategies for staff training, particularly for advanced neonatal care, has emerged as an urgent area of inquiry. Given the relative abundance of neonatologists in high income countries, pairing them as mentors with HCP in low resource settings to facilitate neonatal education is an approach that could help fill this gap. To explore this concept, an email was sent to a list-serve of US based neonatologists, seeking volunteers with global health experience interested in participating in such an effort. Over one hundred respondents requested to participate, enabling this pilot curriculum named “Born To Thrive (BTT).

The content and innovative approach of the BTT curriculum was a first of its kind at KGH. Therefore, we sought to formally assess the effect of the neonatal medicine educational curriculum on improving knowledge of the HCPs at the SL SCBU as evidenced by comparison of pre-test to post-test results, and to understand the perceived value of specific aspects of the educational methodology by the participants, specifically the flipped classroom approach, the mixed classroom learners, and the added value of the in person training compared to the virtual training sessions alone.

## Methods

This was a prospective, interventional, quality improvement, pre-post quasi-experimental study design. Seven US based mentors (four neonatologists, a neonatology fellow, and two neonatal nurse practitioners) were selected from the list of responders, with attention to age, global health experience, race, and gender. After conducting a needs assessment, the US based mentors collaborated with KGH SCBU educational leaders to develop the neonatal BTT curriculum.

The BTT “intervention” was conducted between October 2024 and April 2025. It consisted of two phases of virtual education, each approximately 2 months long, with a one-month break between sessions, followed by a one week in person training focusing on consolidation of knowledge and hands-on skill development. The purpose of the curriculum was to train and support the HCPs working at KGH’s SCBU by facilitating education for prevention, diagnosis, and advanced management of neonatal conditions. Both printed and verbal material were provided in English, which is the language used in the KGH hospital setting.

All KGH SCBU nurses, midwives, MOs, and CHOs (the participants) were invited to take part in the intervention. They completed a total of four educational assessments consisting of a pre-test and post-test at the beginning and end of each phase. They also completed a subjective assessment consisting of Likert Scale and open text questions at the end of the intervention to capture their perception of the teaching methodologies in the curriculum.

Unlike the traditional classroom approach to teaching, which was common in previous trainings at this site, the BTT intervention used an innovative adult-learning based approach with three key attributes:

i. A “flipped classroom” style in which participants watched prerecorded lectures and/or videos that covered the majority of the content, followed later in the week by live virtual sessions in which the US based mentors reinforced the concepts from the prerecorded sessions using cases, interactive scenarios, and question/answer sessions.
ii. A multi-disciplinary classroom with nurses, midwives, MOs, and CHOs training together as a team rather than a single population classroom learning environment.
iii. Two virtual teaching phases followed by an in-person education session to address learning deficiencies identified in the virtual curriculum and focus on hands-on skills such as procedures, resuscitation, and simulation-based teaching.

### Educational Content

KGH SCBU leadership shared the results of their needs assessment regarding gaps in knowledge and skills necessary to provide advanced care to SSNBs. They also provided the Sierra Leone National Neonatal Protocol and a list of available medications and equipment to ensure that the BTT curriculum was contextually adapted and appropriate. During a series of standing weekly meetings, the mentors voluntarily chose topics to develop and teach based on their area of interest, strength and experience. In most cases two mentors shared a topic. The mentors responsible for each topic developed the prerecorded lecture or video material by recording a lecture of themselves covering the topic and/or finding on-line open access content. They then created the material to share for the live virtual session, typically a slide presentation intended to promote discussion and interactions. This material was all shared with the SCBU leadership, who provided edits based on local knowledge. The final versions were then uploaded onto a shared online drive at least a week ahead of schedule. The process was repeated for Phase 2.

The topics covered in Phases 1 and 2 are listed in Fig 1. The content of the Phase 3 in-person visit was based on identified learning deficiencies from Phase 1 and 2 post-test results, as well as input from the SCBU leadership regarding opportunities to strengthen hands on skills: Hyperbilirubinemia and phototherapy, risk factors with small for gestational age/large for gestational age, intraventricular hemorrhage (IVH) reduction best practices, infection control and prevention, routine care of newborn and newborn anthropometry, preparation of the delivery area, Essential Newborn Care (ENC), Objective Structured Clinical Examination (SCE), PIV line care and IV fluid preparation, and delivering difficult news simulation and debrief.

**Figure 1.**
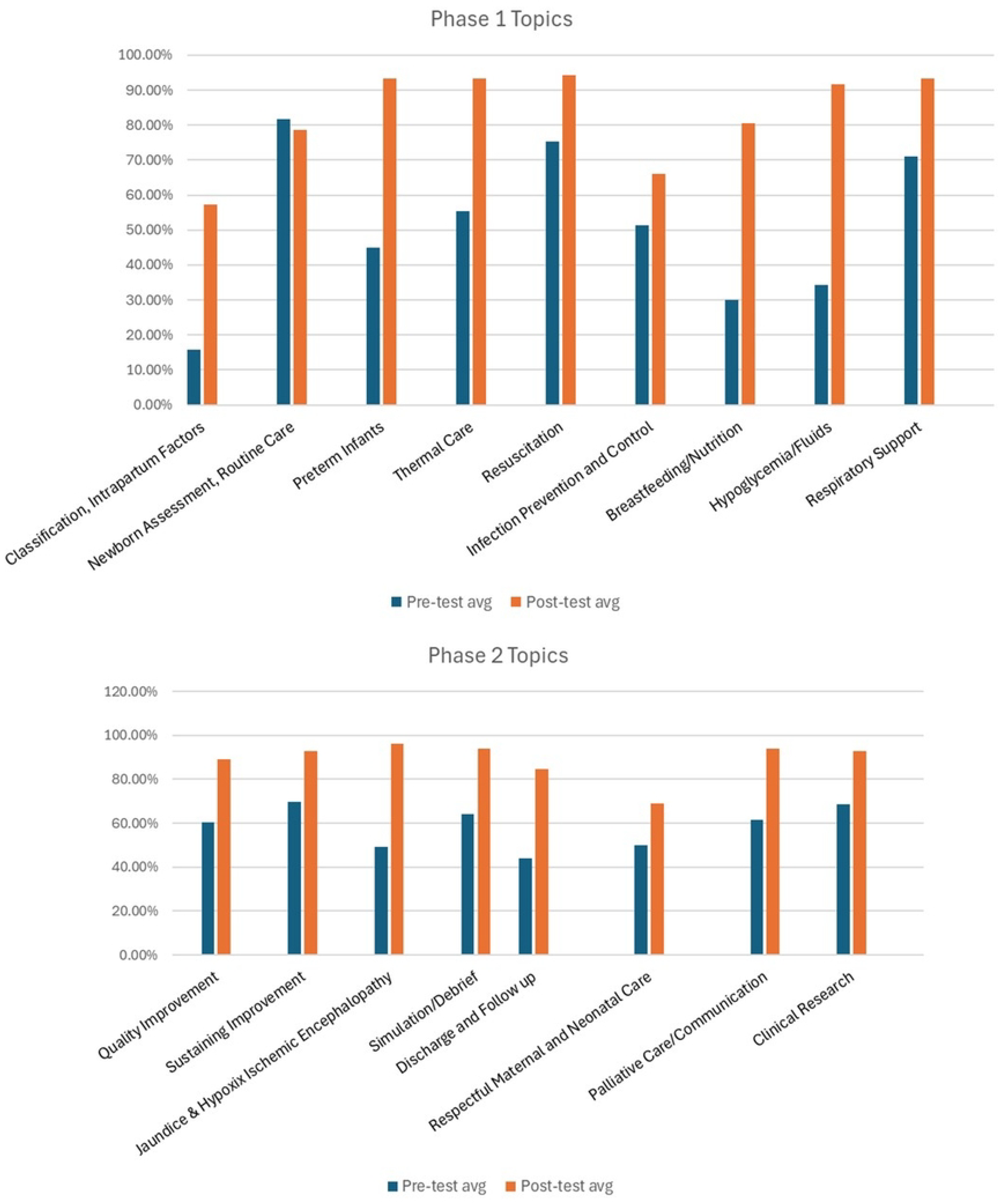
Topics by Phase with Comparison of Average Pre-Test and Post-Test Results.

### Educational Assessments

The pre- and post-test assessments consisted of 27 questions for Phase 1 and 25 questions for Phase 2. The pre-test questions were the same as the post-test questions. Questions were developed by US-based Neonatology mentors assigned to facilitate the topic and were based directly on the learning objectives of the session. Each topic was assessed based on approximately three questions. The pre- and post-tests were completed immediately before and after each phase. The subjective assessment consisted of six Likert Scale questions, one picklist of preferred learning modalities, and seven free text responses. They were completed at the end of the in-person Phase 3 training.

To accommodate the most participants and avoid overlapping learning with clinical duties, one lesson was scheduled for the day shift just after they signed out, and another session scheduled for the night shift just before they signed in. A WhatsApp (WhatsApp, Menlo Park, California, USA) chat group was set up to facilitate communication between all participants, US based mentors, and organizers. This was critical for reminders, and to motivate participants to take the pre-/post-tests and attend the sessions. All participants who attended at least 85% of the teaching sessions were designated as completing the BTT curriculum. The participants celebrated the completion of the course at the end of Phase 3 with a certificate-based ceremony.

We performed standard descriptive statistics on the participants’ characteristics, including such data as hospital role, years since graduation, and gender of participant. Frequencies and percentages were used to describe the characteristics of participants. The median and interquartile range (IQR) were used to summarize the raw pre- and post-test scores in Phase 1 and Phase 2 with the Wilcoxon signed rank test used to test for a pre-versus post-test change. Only participants with both pre- and post-test scores were included in the comparison. The Likert Scale responses to the subjective assessments were reported as percentages. The open text answers were divided into overarching categories and reported as percentages. For the picklist and open text answers, because the categories were not mutually exclusive, an individual’s responses could fall into more than one category. Therefore, the number of responses may exceed the number of participants.

Ethics approval was provided by the Sierra Leone Ethics Review Board and the Partners In Health Sierra Leone Research Department. The hospitals employing each of the affiliated US based mentors were notified of the study and either gave full institutional review board (IRB) approval or provided an exemption. Written consent was obtained from all participants.

## Results

Thirty-eight HCPs participated in the curriculum, of whom 27 took both the pre- and post-tests for Phase 1 and 25 took both the pre- and post-tests for Phase 2. Characteristics of participants are shown in Table 1. Aggregated scores in both Phases improved for all participants who took both the pre- and post-tests (Table 2). Average scores for Phase 1 and Phase 2 increased significantly at post-test compared to pre-test except for the Newborn Assessment and Routine Care topics in Phase 1 (Fig 1).

**Table 1.** Participant Characteristics (n = 38)

| Characteristic | n (%) |
| --- | --- |
| **Gender** |  |
| Female | 25 (65.8%) |
| **Position** |  |
| Nurse (Registered and Community Health Nurse) | 27 (71.1%) |
| Midwife | 3 (7.9%) |
| Medical Officer (MO) | 1 (2.6%) |
| Community Health Officer (CHO) | 6 (15.8%) |
| Maternal and Child Health Aid | 1 (2.6%) |
| **Ward** |  |
| Labor | 2 (5.3%) |
| NICU | 29 (76.3%) |
| Pediatric | 7 (18.4%) |
| **Length of Experience** |  |
| 1 - <2 years | 4 (10.5%) |
| 2 - <3 years | 9 (23.7%) |
| 3 - <4 years | 5 (13.2%) |
| 4 - <5 years | 10 (26.3%) |
| ≥ 5 years | 10 (26.3%) |

**Table 2.** Aggregated Scores for Pre- and Post-Tests by Phase (Complete Pre-Post Pairs Only)

| Phase | Pre-Test Median (IQR) | Post-Test Median (IQR) | P-value |
| --- | --- | --- | --- |
| Phase 1 (n = 27) | 14.0 (12.0, 16.0) | 23.0 (22.0, 23.0) | < 0.001 |
| Phase 2 (n = 25) | 14.0 (11.8, 17.0) | 23.0 (21.8, 23.0) | < 0.001 |

*Only participants with both pre-test and post-test scores are included. IQR = interquartile range (Q1, Q3). P-values from Wilcoxon signed-rank test (paired, two-sided).*

Thirty-one participants completed the subjective assessment at the conclusion of the curriculum (Table 3). On the Likert scale section 97% (30/31) rated the curriculum to be “very effective”. Most participants (24/31, 77%) found the pre-recorded materials to be “very effective,” and (25/31, 81%) found the live virtual sessions were “very effective.” Most participants (27/31, 87%) found that the prerecorded materials prepared them well for the live virtual sessions. All participants (31/31, 100%) indicated that the week of in-person instruction provided at the end of the curriculum was “very helpful.” All participants (31/31, 100%) reported that they liked or strongly liked learning in a multidisciplinary group (nurses, midwives, MOs, and CHOs).

**Table 3.**
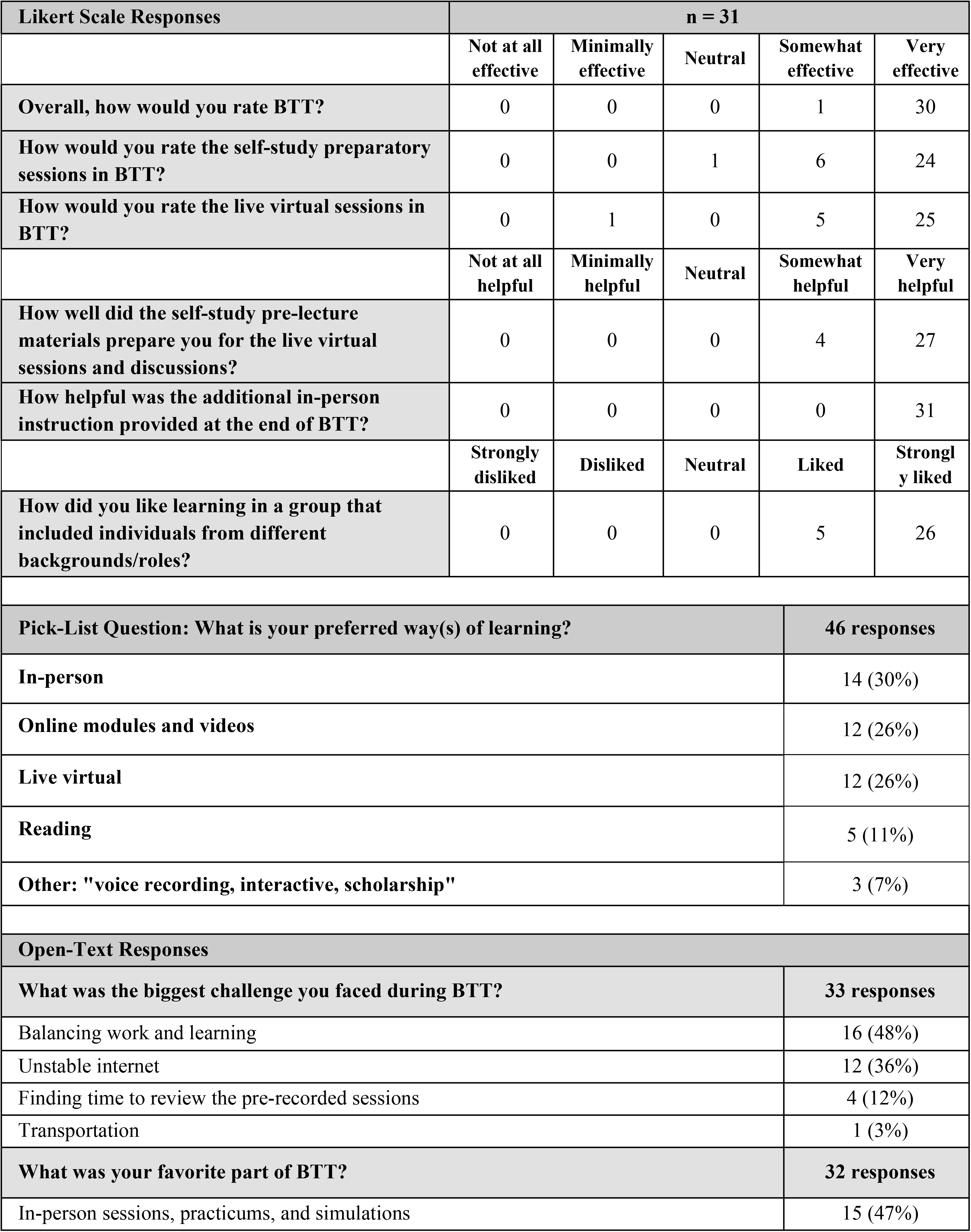
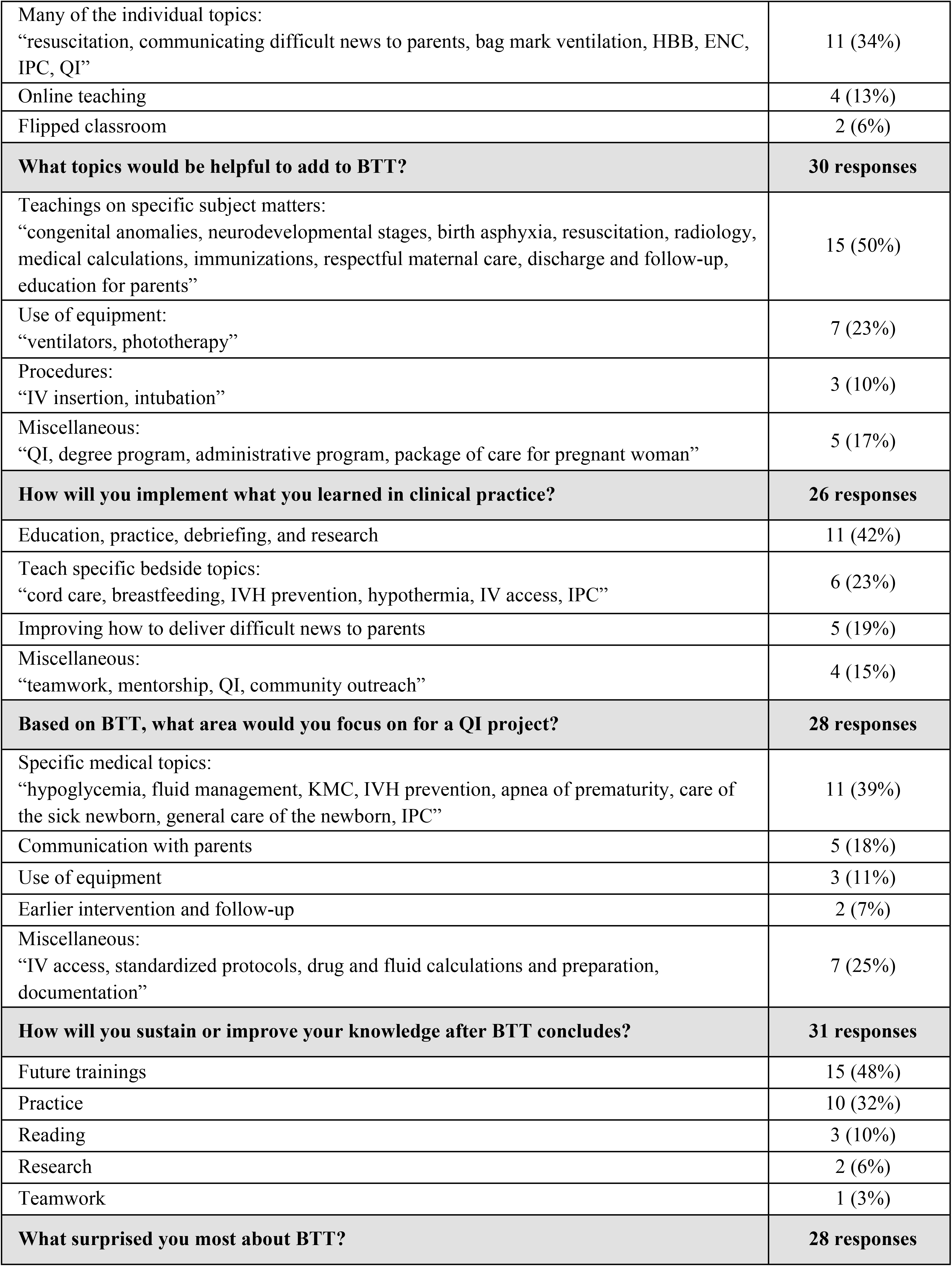
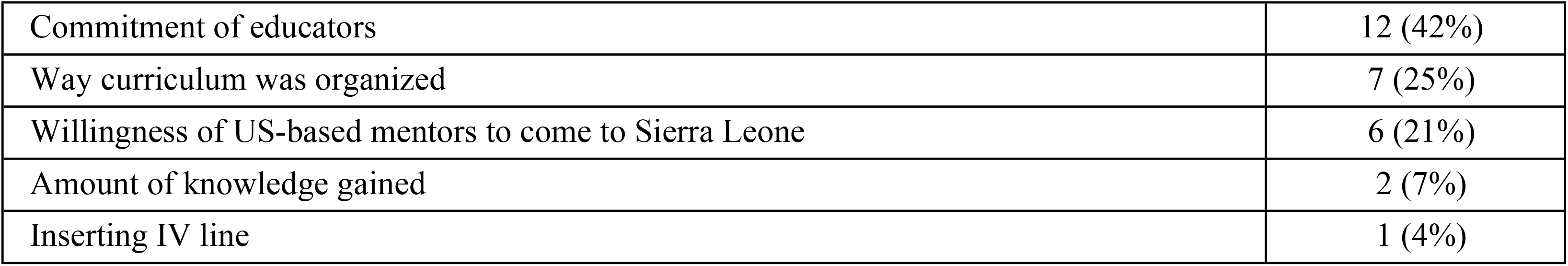
Subjective Assessment.

| Likert Scale Responses | n = 31 |  |  |  |  |
| --- | --- | --- | --- | --- | --- |
|  | Not at all effective | Minimally effective | Neutral | Somewhat effective | Very effective |
| Overall, how would you rate BTT? | 0 | 0 | 0 | 1 | 30 |
| How would you rate the self-study preparatory sessions in BTT? | 0 | 0 | 1 | 6 | 24 |
| How would you rate the live virtual sessions in BTT? | 0 | 1 | 0 | 5 | 25 |
|  | Not at all helpful | Minimally helpful | Neutral | Somewhat helpful | Very helpful |
| How well did the self-study pre-lecture materials prepare you for the live virtual sessions and discussions? | 0 | 0 | 0 | 4 | 27 |
| How helpful was the additional in-person instruction provided at the end of BTT? | 0 | 0 | 0 | 0 | 31 |
|  | Strongly disliked | Disliked | Neutral | Liked | Strongly liked |
| How did you like learning in a group that included individuals from different backgrounds/roles? | 0 | 0 | 0 | 5 | 26 |
| Pick-List Question: What is your preferred way(s) of learning? |  |  |  |  | 46 responses |
| In-person |  |  |  |  | 14 (30%) |
| Online modules and videos |  |  |  |  | 12 (26%) |
| Live virtual |  |  |  |  | 12 (26%) |
| Reading |  |  |  |  | 5 (11%) |
| Other: "voice recording, interactive, scholarship" |  |  |  |  | 3 (7%) |
| Open-Text Responses |  |  |  |  |  |
| What was the biggest challenge you faced during BTT? |  |  |  |  | 33 responses |
| Balancing work and learning |  |  |  |  | 16 (48%) |
| Unstable internet |  |  |  |  | 12 (36%) |
| Finding time to review the pre-recorded sessions |  |  |  |  | 4 (12%) |
| Transportation |  |  |  |  | 1 (3%) |
| What was your favorite part of BTT? |  |  |  |  | 32 responses |
| In-person sessions, practicums, and simulations |  |  |  |  | 15 (47%) |

|  |  |
| --- | --- |
| Many of the individual topics:<br>“resuscitation, communicating difficult news to parents, bag mask ventilation, HBB, ENC, IPC, QI” | 11 (34%) |
| Online teaching | 4 (13%) |
| Flipped classroom | 2 (6%) |
| **What topics would be helpful to add to BTT?** | **30 responses** |
| Teachings on specific subject matters:<br>“congenital anomalies, neurodevelopmental stages, birth asphyxia, resuscitation, radiology, medical calculations, immunizations, respectful maternal care, discharge and follow-up, education for parents” | 15 (50%) |
| Use of equipment:<br>“ventilators, phototherapy” | 7 (23%) |
| Procedures:<br>“IV insertion, intubation” | 3 (10%) |
| Miscellaneous:<br>“QI, degree program, administrative program, package of care for pregnant woman” | 5 (17%) |
| **How will you implement what you learned in clinical practice?** | **26 responses** |
| Education, practice, debriefing, and research | 11 (42%) |
| Teach specific bedside topics:<br>“cord care, breastfeeding, IVH prevention, hypothermia, IV access, IPC” | 6 (23%) |
| Improving how to deliver difficult news to parents | 5 (19%) |
| Miscellaneous:<br>“teamwork, mentorship, QI, community outreach” | 4 (15%) |
| **Based on BTT, what area would you focus on for a QI project?** | **28 responses** |
| Specific medical topics:<br>“hypoglycemia, fluid management, KMC, IVH prevention, apnea of prematurity, care of the sick newborn, general care of the newborn, IPC” | 11 (39%) |
| Communication with parents | 5 (18%) |
| Use of equipment | 3 (11%) |
| Earlier intervention and follow-up | 2 (7%) |
| Miscellaneous:<br>“IV access, standardized protocols, drug and fluid calculations and preparation, documentation” | 7 (25%) |
| **How will you sustain or improve your knowledge after BTT concludes?** | **31 responses** |
| Future trainings | 15 (48%) |
| Practice | 10 (32%) |
| Reading | 3 (10%) |
| Research | 2 (6%) |
| Teamwork | 1 (3%) |
| **What surprised you most about BTT?** | **28 responses** |
| Commitment of educators | 12 (42%) |
| Way curriculum was organized | 7 (25%) |
| Willingness of US-based mentors to come to Sierra Leone | 6 (21%) |
| Amount of knowledge gained | 2 (7%) |
| Inserting IV line | 1 (4%) |

For the picklist, the majority of participants preferred in-person education (14/31, 45%), or live virtual education sessions (11/31, 35%) compared to prerecorded modules (9/31, 29%) or self-reading (5/31, 16%).

The free text responses provided information on challenges, strengths and next steps (Table 3). Participants’ biggest challenges were balancing working and learning (48%); unstable internet (36%); and finding time to review the prerecorded sessions (12%). Their favorite part of the course included the in-person sessions, practicums and simulations (47%); and many of the individual topics (34%), especially neonatal resuscitation and communicating difficult news to parents. When asked for helpful additional topics, most requested teachings on specific subject matters (50%) such as congenital anomalies, neurodevelopmental stages, and medication calculation; and use of equipment (23%) such as ventilators and phototherapy. In terms of how they would implement what they learned in their clinical practice, they thought they could facilitate ongoing education, practice, debriefing, and research (42%); bedside teaching of specific topics (23%) such as cord care, breast feeding, prevention of IVH and hypothermia; and improve how they gave difficult news to parents (19%). When asked which area they would choose for a quality improvement (QI) project, they had a wide range of ideas including specific medical topics (39%) such as hypoglycemia, fluid management, Kangaroo Mother Care, and communication with parents (18%); and use of equipment (11%). To sustain or improve their knowledge after the course was completed, most recommended future trainings (48%) and practice (32%) but some also recommended reading, teamwork and research. Finally, what surprised them most about the curriculum was the commitment of the educators (42%), the way the curriculum was organized (25%), and the willingness of the US-based mentors to come to Sierra Leone (21%).

## Discussion

To meet the recommendations of WHO’s recommendations for SSNBs, Sierra Leone requires adequate numbers of skilled newborn HCPs.^3^ This requires adequate numbers and training of HCPs.^6^ As care levels increase from basic neonatal essentials to tertiary neonatal intensive care, the knowledge and skill level of the nurses, midwives, MO and CHOs need to increase in parallel. Despite this, in low-income countries, HCP can be assigned to a wide range of illness severity on neonatal units, working independently after no to minimal orientation.^7^ In most low-income countries including Sierra Leone, nurses, midwives, MOs and CHOs without speciality training are called to provide care for SSNBs regardless of the level of care required.

While the need for neonatal training in low-resource settings is clear, the variations in medications, equipment, and baseline staff training make a common curriculum challenging. Multiple individual studies and meta-analyses have shown that teaching the WHO ENC and Helping Babies Breathe (HBB) curricula improve perinatal outcomes such as stillbirth rate, very early neonatal mortality, and overall neonatal mortality.^8,9,10,11,12^ However, there currently is not an established training program for more advanced neonatal care in LMICs. Thus, the need for a culturally-adapted, contextually-appropriate training program on improving the care of SSNBs beyond foundational principles within the ENC and HBB frameworks is needed.

Several studies have demonstrated the feasibility and effectiveness of virtual education. Virtual training of neonatal resuscitation skills in LMICs have also been shown to be feasible and may be cost-effective.^13,14,15,16,17,18,19,20,21^ Thus, as the deadline of the SDGs rapidly approaches, innovative and effective strategies for staff training, particularly for advanced neonatal care, emerge as an urgent area of inquiry.

Our BTT curriculum builds on virtual education, by using flipped classroom methods for virtual twice weekly discussions, and a highly interactive in-person education and skills training to cement the learning in key areas including resuscitation, procedural skills, debriefing, and communication. Overall, we found that this curriculum significantly improved the knowledge base for both phases of the curriculum and was well-regarded by the participants in the SCBU at KGH in Sierra Leone. Specifically, all modules except one (Routine Care and Newborn Assessment) demonstrated improved knowledge scores when comparing the pre-to post-test scores. Notably, the Routine Care and Newborn Assessment module had the highest pre-test score of all domains, and the scores were not significantly different between pre- and post-tests. This suggests that, due to the high pre-training knowledge demonstrated in this module, the content may need to be adapted to enhance new knowledge acquisition such as teaching more advanced assessment skills and adding information on more advanced topics such as congenital anomalies.

The areas with the most robust improvement in knowledge from pre-to post-test included: neonatal nutrition and optimizing breastfeeding practices, identification and treatment of neonatal hypoglycemia, intravenous fluid prescription and delivery, prenatal factors affecting neonatal outcomes, classification of neonates based on anthropometric assessments, identification and treatment of neonates with hypoxic ischemic encephalopathy, and discharge and follow up practices. Areas of high pre-training knowledge included neonatal routine care, neonatal resuscitation, and respiratory support (predominantly related to continuous positive airway pressure [CPAP]). These generally reflect topics already covered by HBB and ENC, and may reflect modules that could be removed in contexts where HBB and ENC are already being implemented. Overall, these findings collectively demonstrate a robust increase in knowledge acquisition across multiple domains by HCPs in Sierra Leone. They further demonstrate the ability of this BTT curriculum to be tailored and contextually adapted into other contexts worldwide where resources and use of other neonatal training modules (e.g. HBB, ENC) may or may not be included. Our study has several challenges and limitations. First, systems-level challenges contributed to the inability to collect reliable clinical outcomes data, limiting our ability to directly link the training with clinical outcome changes. Additionally, lack of funding was a notable challenge; the US-based mentors served in a volunteer capacity and self-funded international travel for the in-person phase. Furthermore, as the course was conducted in English, which is the language within the hospital infrastructure at this hospital as well as the preferred language by the US-based mentors, this will not be applicable to all contexts worldwide. As such, the generalizability may be limited and requires contextual and linguistic adaptations in other settings. Finally, the sustainability of the course using US-based mentors is a concern. As such, the team has developed recordings to assist with future sustainability of this curriculum along with the empowerment of local site-specific champions.

Because of the multiple competing demands on an in-country team, it can be burdensome to perform, analyse and publish the results of a pilot course such as ours. This likely explains the relative paucity of literature on collaborative educational efforts in low-resource settings. The Sierra Leone SCBU team was eager to collaborate with the US-based mentors to create a study design, usher it through multiple IRB processes, and analyse the pre- and post-tests as well as the subjective assessments. This academic aspiration expanded the opportunities for mentorship and capacity building.

Despite the many challenges and limitations, this curriculum had many strengths and successes. First, we conducted a needs assessment prior to the development of the course. This directly informed the content development and enabled the curriculum to be based on contextual, real-world, contemporary concerns. Additionally, the course benefited by having a strong local champion in a nurse educator role as well as the support of hospital administration. The learners were all very engaged, and the use of a WhatsApp chat group, which is a common practice for communication in this setting, enhanced our communication in an approachable, timely, and contextually appropriate manner. Overall, this curriculum was successful in advancing the knowledge of care providers in the SCBU at KGU in Sierra Leone. The participants received the course favourably. The US-based mentors expressed their enthusiasm by uniformly volunteering to adapt the curriculum and teach it at other sites. Ideally, neonatal training would be taught by local educators to improve sustainability. But given the urgent need to expand the knowledge of hospital level neonatal care, our approach represents one low cost, feasible option. The more than 100 remaining US-based mentors who wanted to volunteer for this opportunity represent a large, untapped pool of educators.

## Conclusion

The virtual, flipped classroom, neonatal medicine curriculum that was contextually and culturally adapted was well-regarded. The implementation of this curriculum significantly improved the knowledge base and skill development of SCBU HCPs. Evaluation of the impact of this BTT curriculum on improving neonatal clinical outcomes is a critical next step. Adapting this virtual flipped classroom curriculum or similar curricula show promise to improve the quality of care for SSNBs in low-resource settings.

## Data Availability

The minimal data set is available at Harvard Dataverse via: https://doi.org/10.7910/DVN/RSY2UA

## Acknowledgements

We would like to acknowledge the contribution of Rosemary Bain for her coordination of the Born To Thrive curriculum and this manuscript.

